# Rare splice and missense variants with evidence of pathogenicity in consanguineous families with autosomal recessive intellectual disability from Pakistan

**DOI:** 10.1101/2024.01.08.23299914

**Authors:** Abdul Waheed, Robert Eveleigh, Danielle Perley, Janick St-Cyr, François Lefebvre, Abdul Hameed Khan, Zarqash Majeed, Abrish Majeed, Katerina Trajanoska, Raquel Cuella-Martin, Claude Bhérer, Ghazanfar Ali, Vincent Mooser, Daniel Taliun

## Abstract

Intellectual disability (ID) is a neurodevelopmental disorder affecting up to 1-3% of people worldwide. Genetic factors, including rare *de novo* or rare homozygous mutations, explain many cases of autosomal dominant or recessive forms of ID. ID is clinically and genetically heterogeneous, with hundreds of genes associated with it. In this study, we performed high-depth whole-genome sequencing of twenty individuals from five consanguineous families from Pakistan, with nine individuals affected by mild or severe ID. We identified one splice and five missense rare variants (at allele frequencies below 0.001%) in a homozygous state in the affected individuals with supporting and moderate evidence of pathogenicity based on guidance from the American College of Medical Genetics and Genomics. These six variants mapped to different genes (*SRD5A3*, *RDH11*, *RTF2*, *PCDHA2*, *ADAMTS17*, and *TRPC3*), and only *SRD5A3* had previously been known to cause ID. The p.Tyr169Cys mutation inside *SRD5A3* was predicted to be deleterious and affect protein structure by multiple *in silico* tools. In addition, we found one missense mutation, p.Pro1505Ser, inside *UNC13B* with conflicting evidence of pathogenic and benign effects. Further functional studies are required to confirm the pathogenicity of these variants and understand their role in ID. Our findings provide additional needed information for interpreting rare variants in the genetic testing of ID.

## 1. INTRODUCTION

Studies estimate the prevalence of ID in the general population to be from less than 1% and up to 3%, depending on the country [1–3]. The main characteristics of ID are decreased mental abilities (such as reasoning, recognition, abstract thinking, and judgement) and, consequently, decreased adaptive functioning (such as communication, social participation, and independent living) [3]. ID belongs to a group of neurodevelopmental disorders that typically manifest in early childhood and can co-occur with other neurodevelopmental disorders (e.g., autism spectrum disorder, ASD) or physical conditions (e.g., cerebral palsy). Depending on the severity level (mild, moderate, severe, profound), ID may be diagnosed within the first two years of life (severe) or may not be identifiable until school age (mild) [3]. Early and accurate ID diagnosis is essential for initiating early intervention strategies to improve the quality of life of those affected.

Both genetic and non-genetic factors can cause ID. Non-genetic factors include events and environmental exposures during and after pregnancy (e.g., maternal disease, exposure to hazardous chemicals) and early childhood (e.g., severe head injury, meningitis, chronic social deprivation). Genetic factors consist of chromosomal aneuploidies and rare mutations, including copy-number variations (CNVs), single-nucleotide variations (SNVs), and structural variations (SVs). Rare *de novo* mutations may lead to autosomal dominant ID (ADID) and are estimated to explain up to 60% of severe ID cases [4]. Rare mutations inherited in the homozygous state by individuals, typically in consanguineous families, are associated with autosomal recessive ID (ARID). They are estimated to explain up to 57% of ARID cases in consanguineous families [4]. ID is a genetically heterogeneous disease with up to 700 genes linked to it or associated disorders by 2015 [4], and around 1,300 genes listed in the approved gene panel for ID (v5.0 from 22 March 2023) available at Genomics England’s PanelApp [5]. However, the prevailing view is that more ID-causal genes are yet to be discovered, and the effects of all possible mutations within ID-causing genes have not yet been assessed [4, 6, 7]. Searching and describing novel rare mutations inside known ID genes and identifying new candidate genes may improve the molecular diagnosis of ID and lead to a better understanding of the underlying molecular disease mechanisms.

This study focuses on identifying rare mutations responsible for ARID in five consanguineous families from Azad Jammu and Kashmir, Pakistan. We performed WGS in twenty family members (nine affected by mild, moderate, or severe ID) with a projected average depth of coverage of 30X. We called SNVs and short insertions and deletions (indels) and followed them with extensive quality checks, variant filtering, and *in silico* variant effect predictions. Then, we performed mapping of autosomal recessive variants for ID in each family using only high-quality, rare and predicted potentially damaging variants. In this stage, we limited our analysis to rare variants within protein-coding gene boundaries, which include coding and untranslated regions (UTRs) and introns. We identified seven mutations in homozygous state in affected family members placed within seven genes (*SRD5A3*, *RDH11*, *RTF2*, *PCDHA2*, *ADAMTS17*, *UNC13B*, and *TRPC3*), where only *SRD5A3* was established previously as causal of ID. Each of the seven mutations showed one moderate and one supportive evidence of pathogenicity as defined in the American College of Medical Genetics and Genomics (ACMG) guidance for the interpretation of sequence variants.

The first section of the results describes the variant filtering and quality control across all sequenced individuals. We then describe the findings in each family separately. The last section describes the results of *in silico* modelling of protein structures with the identified possibly damaging variants.

## 2. METHODS

### 2.1. Study description

Five consanguineous families with Autosomal Recessive Intellectual Disability (ARID) were identified in various urban and rural areas of Azad Jammu and Kashmir, Pakistan (Supplementary Figure 1). Only families including two or more persons suffering from concordant phenotypes related to ARID, having parental consanguinity and showing the autosomal recessive mode of inheritance, were enrolled in the study. For simplicity, analyzed families are referred to as ARID-76, ARID-78, ARID-79, ARID-80, and ARID-81. The ORIC (Office of Research Innovation and Commercialization) and DSAR (Director of Advanced Study and Research) boards of the University of Azad Jammu and Kashmir, Muzaffarabad, Pakistan, approved the study ethics protocol. Written informed consent forms were signed by the guardians of each family enrolled in current research for performing genetic analyses and publishing face photographs whenever necessary. The DNA sequencing and genetic data analyses were also approved by the Research Ethics Office (IRB) of the Faculty of Medicine and Health Sciences at McGill University, Montreal, Canada.

### 2.2. Interviews and clinical evaluation

Participating families were interviewed in person. Parents or guardians were interviewed regarding the family history of the affected persons to rule out the involvement of any environmental factors. No environmental factors were identified. A trained physician conducted a physical examination during a home visit to exclude additional morphological, behavioural, neurological, or other organ system abnormalities. Affected individuals were assessed for the presence of ID (mild, moderate, or severe), developmental delay, deficient behaviour, and other co-morbid phenotypes (e.g., communication, behaviour with others, aggressiveness, anxiety, sleeping time, awareness of daily life needs, locomotion, and movements). From three-to five-generation pedigrees were collected and analyzed to infer the mode of inheritance of the disease. Based on the information provided, pedigrees were structured following the approach described in Bennett *et al*., 2008 [8]. Genetic relationships among individuals and disease inheritance patterns prevailing within the families have been elucidated through pedigrees. Each family pedigree was anonymized and assigned a unique identifier consisting of the ARID-prefix and a two-digit number, and the backward mapping is known only within the research group.

### 2.3. Blood sample collection and DNA extraction

Venous blood was collected from affected and unaffected family members. Blood was sampled and preserved in vacutainer tubes containing ethylene diamine tetra acetate (EDTA). Blood samples were transported from the field in the ice box. Samples were stored in the refrigerator at the Department of Biotechnology University of Azad Jammu and Kashmir lab. Genomic DNA was extracted from peripheral white blood cells using the standard phenol-chloroform method.

### 2.4. Sequencing, alignment, and quality checks

Whole genome sequencing (WGS) was performed at the McGill Genome Centre in October 2022, targeting 300 bp length fragments and 30X depth of sequencing coverage (DP). Raw reads sequencing data was aligned to the GRCh38 reference genome, sorted by position, indels re-aligned, marked for duplicates and quality controlled using GenPipes version 4.3.0 [9]. The aligned reads were stored in BAM files. The average DP of WGS across individuals was 27.82X (SE =1.92X) (Supplementary Table 1). On average, 90.52% (SE=0.26%) of the sequenced base pairs were covered by more than ten high-quality reads (i.e., DP > 10X). There were no differences in average DP and percent of base pairs with DP>10X between affected and unaffected individuals and no individuals with significantly lower average DP. (Supplementary Figure 5). The average Freemix contamination score estimated using VerifyBamId2 [10] was 3.33×10^-4^ (SE = 2.26×10^-4^), suggesting no evidence of DNA contamination. The estimated number of copies of sex chromosomes derived from the sequencing data was consistent with the reported sex phenotype (Supplementary Figure 6). Principal component analysis (PCA) of the sequenced data showed that all individuals had Central South Asian genetic ancestry, consistent with the known demographic history of the local population (Supplementary Figure 7).

### 2.5. Variant calling and quality checks

GATK’s HaplotypeCaller v4.3.0 [11] was used for variant calling. Variant calls were generated in the standard variant calling files (VCFs) – one VCF per individual. Indels were left-aligned using “bcftools norm” v1.13 [12] to generate normalized VCFs. We used three filters to identify and exclude low-quality variants: GATK CNN1D (convolutional neural network, 1D model), GATK CNN2D (convolutional neural network, 2D model), and GATK Hard Filters with GATK’s recommended thresholds (Supplementary Figure 2). Then, we used consensus filtering to keep only those variants that passed all the abovementioned filters (Supplementary Figure 3). We processed the individual BAM files to compute the depth of coverage at each chromosomal position for each sample. Then, we aggregated the depth of coverage at each chromosomal position across all samples and generated an accessibility mask – a list of regions where all individuals were sequenced with 10X or higher depth of coverage. Using the generated accessibility mask, we subset variants for each individual to create a final per-individual VCF, which includes only variants overlapping regions in the accessibility mask. Finally, we merged individual VCFs into a single VCF. We kept only those variants which passed GATK filters across all individuals. When an individual did not have an alternate allele at a specific position within the accessibility mask, the genotype was set to 0/0. The total number of SNVs and indels is reported in Supplementary Table 2.

### 2.6. Mapping of autosomal recessive variants for ID

We annotated genetic variants with the Variant Effect Predictor (VEP) tool (release 109) [13]. To map autosomal recessive variants for ID, we considered only variants which are missense, nonsense (stop gain, stop lost, start lost), splice site (splice donors, splice acceptors), frameshifts, and are within 5’ and 3’ UTRs. We kept only those variants which were predicted as potentially damaging by at least one method: PolyPhen > 0.15, SIFT < 0.05 or CADD (Phred-scaled) > 20. We considered two alternate allele frequency (AF) thresholds for selecting rare variants based on gnomAD v3.1.2 [14] and 1000 Genomes Project Phase 3 [15] datasets (i.e., AF must be less than the selected threshold in both datasets): AF < 0.01% and AF < 0.001% (Supplementary Figure 4). For the selected predicted potentially damaging rare variants, we looked at their states in each ARID family. We kept only those which are homozygous (i.e., genotype 1/1) in affected members of the family, heterozygous (i.e., genotype 1/0) in non-affected parents; and heterozygous or absent (i.e., genotype 0/0) in non-affected siblings. For the variants that passed the filters mentioned above, we looked up whether they were previously reported in the ClinVar database (accessed in November 2023) [16].

### 2.7. Conservation

The conservation of the ancestral allele across species was assessed with phyloP and phastCons conservation scores [17–21] computed based on the multiple alignments of 100 vertebrates from the UCSC Genome Browser GRCh38/hg38 release [22] or GERP conservation scores [23] calculated based on the multiple alignments of 91 mammals from the Ensembl genome browser release 110 [24]. The phyloP method assesses conservation at individual nucleotides, ignoring the effects of their neighbours, and calculates -log(*P*-value) under a null hypothesis of neutral evolution. Positive phyloP scores correspond to conservation. The phastCons method computes the probability that a nucleotide belongs to a conserved element, i.e., it considers the neighbouring nucleotides. Similarly to phyloP, the GERP method measures conservation at individual nucleotides and compares the number of observed substitutions to the expected under no functional constraint. Positive GERP scores represent conservation. Our analyses classified the ancestral allele as conserved if at least one of the following thresholds was satisfied: phyloP > 1.9, phastCons > 0.5, GERP > 2.7 [25]. Additionally, we visually inspected the presence of the ancestral allele across multiple alignments of ten primates in the Ensembl genome browser release 110.

### 2.8. Gene constraints

We used the following gene constraint metrics available from gnomAD v2.1.1 and v4.0.0 [26]: the ratio of the observed to expected (o/e) number of loss-of-function (LoF) variants in a gene – values close to 0 indicate that the gene is intolerant to LoF mutations (the recommended threshold for the LoF intolerant gene is when the 90% CI’s upper bound of o/e LoF [LOEUF] is less than 0.35 in gnomAD v2.1.1 and less than 0.6 in gnomAD v4.0.0); the o/e number of missense variants in a gene – values close to 0 indicate that gene is intolerant to missense mutations; missense Z-score – high scores indicate that gene is intolerant to missense mutations (the recommended threshold for the missense intolerant gene is when Z-score > 3.09 [27]). We also used the sub-genic missense constraint score [28], denoted γ, available from the DECIPHER online tool [29]. The γ score varies between 0 and 1, where smaller values indicate fewer observed missense variants than expected.

### 2.9. Prediction of protein stability

We applied four methods to evaluate the effects of the mutations on protein structures: FoldX[30], Rosetta[31, 32], Venus[33], and Missense3D[34]. We looked for the experimentally derived protein structures (e.g., using cryo-electron microscopy (cryo-EM) or X-ray crystallography) from the Protein Data Bank in Europe (PDBe) [35] and for structural predictions from the AlphaFold Protein Structure Database (AlphaFold DB) [36]. The FoldX, Rosetta, and Venus methods predict the change in Gibbs free energy, denoted as ΔΔG and measured in kcal/mol units, between wild-type and mutant protein structures. Mutations resulting in ΔΔG < -1 are considered to stabilize the protein, -1 < ΔΔG < 1 - to be neutral, and ΔΔG > 1 - to destabilize the protein [37]. We ran FoldX v5 locally using default settings: 1) we minimized the original protein structures with the RepairPDB command before estimating ΔΔG; 2) for each mutation, we estimated ΔΔG using the BuildModel command with five runs and calculated the mean ΔΔG value. We ran Rosetta r351 locally following the Cartesian ΔΔG protocol [38]: 1) we used FastRelax[39–41] with the ref2015_cart weights[38] to minimize the original protein structure; 2) for each mutation, we estimated ΔΔG using the cartesian_ddg application [37] with options described in Frenz et al. [38] and five iterations; 3) we applied the 1/2.94 scaling factor to the average ΔΔG [37]. We used the MichelANGlo web application [42] to run Venus, which uses PyRosetta[43] and the speed-optimized protocol based on the REF15 score function [44] and provides scaled ΔΔG estimates. We ran Missense3D through the Missense3D web application [34] with default settings and by uploading the protein structures downloaded from PDBe (if available) or AlphaFoldDB. The Missense3D pipeline evaluates the impact of a mutation on 17 features related to protein structure and highlights features most likely altered by the mutation.

## 3. RESULTS

### 3.1. Variant discovery, quality control and filtering

We identified 4,031,141 (SE=13,231) SNVs and 928,353 (SE=8,962) indels on average per individual (Supplementary Table 2). We didn’t observe systematic differences in the number of identified variants between affected and unaffected individuals (Supplementary Figure 8). After applying variant-level filtering, the average number of SNVs and indels per individual were 3,712,456 (SE=13,013) and 880,956 (SE=7,965), respectively.

In total, we identified 8,325,148 high-quality, unique genetic variants across all individuals placed within genomic regions where each individual had more than ten high-quality reads (i.e., DP>10X) (Supplementary Table 3). We further limited our search to 155,658 non-synonymous variants inside protein-coding regions, splicing regions, and 5’ and 3’ UTRs. Of these, 11,341 variants were predicted as potentially damaging by at least one of the three bioinformatic tools: PolyPhen, SIFT, and CADD. Among these predicted potentially damaging variants, 3,049 had AF < 0.01% in the gnomAD v3.1.2 and 1000 Genomes Project Phase 3 variant databases, with 1,734 remaining when applying a more stringent AF < 0.001% threshold.

### 3.2. The p.Tyr169Cys mutation inside the known *SRD5A3* gene in the ARID-76 family

ARID-76 is a five-generation pedigree in which consanguineous parents have three affected and two unaffected children (Figure 1). WGS was done on three individuals: IV-1 (unaffected parent), V-1 (unaffected sibling) and V-4 (affected sibling). The affected sibling (V-4) shows moderate intellectual impairment with night blindness. They express themselves through non-verbal communication and have limited facial recognition skills. They require help with self-care activities. The second affected sibling (not sequenced) is more severely affected, completely non-verbal, with impaired facial recognition capabilities.

**Figure 1.**
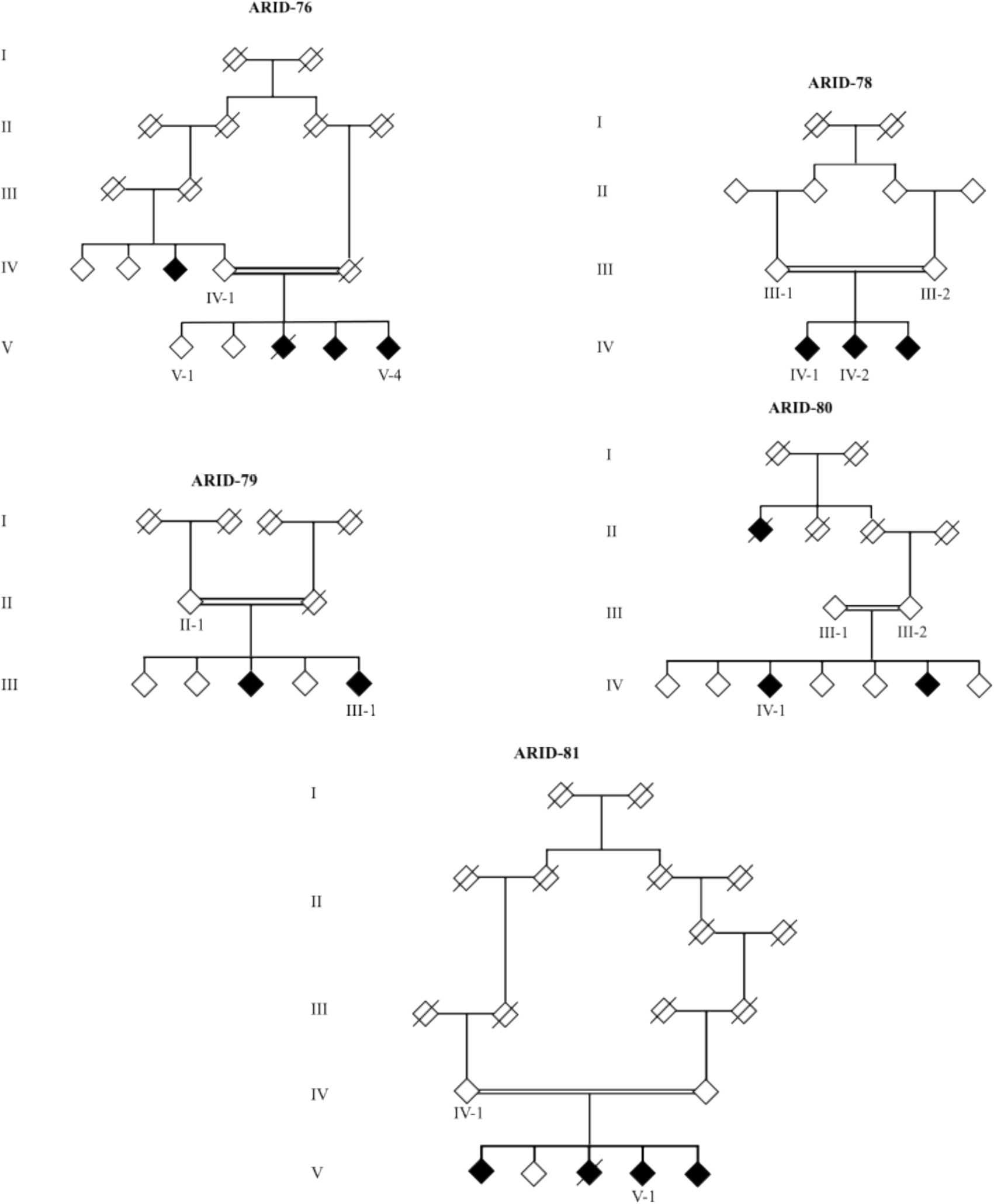
Anonymized partial family trees. Symbols represent family members. Parallel dual lines indicate consanguineous marriages. Shaded and empty symbols represent affected and unaffected family members, respectively. Slanted crossed lines represent deceased family members. Every generation within a pedigree is referred to by Roman numerals (I, II, III, IV) from top to bottom, while the positions of family members are depicted using Arabic numbers (1, 2, 3, 4). WGS was performed on labeled individuals: ARID-76 family – IV-1, V-1 and V-4; ARID-78 family – III-1, III-2, IV-1, and IV-2; ARID-79 family – II-1 and III-1; ARID-80 family – III-1, III-2, and IV-1; ARID-81 family – IV-1 and V-1.

We found a single predicted potentially damaging variant in the homozygous state in the sequenced affected child but not in the sequenced unaffected relatives (Table 1). The affected individual carries two copies of the alternate allele G of the missense variant rs754837473 A>G (p.Tyr169Cys) in the *SRD5A3* gene. In contrast, the unaffected parent has only one copy, and the unaffected sibling has no copies of this allele. The alternate allele G was previously reported by two submitters in the ClinVar database (accession ID: VCV002506565.2) as having uncertain significance [45]. Only one submitter provided a description, although very brief, of a patient condition: the alternated allele G was found in a homozygous state in a young (< 9-year-old) female of South-East Asian ancestry with *SRD5A3*-congenital disorder of glycosylation (SRD5A3-CDG, MIM: 612379), showing a global developmental delay with nystagmus and characterized as self-absorbed, having a happy demeanour, and hand-flapping. In our analyses, the variant was predicted to be potentially damaging by all three *in silico* methods, Polyphen, SIFT, and CADD (Table 2). The comparative genomic analyses suggested that the ancestral allele A is conserved (PhyloP = 9.1, PhastCons = 1.0), and it was present across ten primates (Supplementary Table 4). Only a single copy of the alternate allele G was found in the gnomAD v3.1.2 non-neuro database (AF=6.6×10^-6^), which excludes cases with neurological or psychiatric diseases, and it was reported in an individual of European genetic ancestry. Twelve copies of allele G were found in the gnomAD v4.0.0 database (AF=7.4×10^-6^), which includes 416,555 exomes from the UK Biobank [46], and all observed copies were in the heterozygous state. Furthermore, 8 of 12 reported carriers were of South Asian genetic ancestry, suggesting a higher prevalence of allele G in this population. Although, in general, the *SRD5A3* gene tolerates LoF and missense mutations (Supplementary Table 5), nonsense and frameshift mutations in homozygous and compound heterozygous states in this gene have been implicated in two genetic disorders related to ID. One is SRD5A3-CDG (MIM: 612379), which may result in ID, ophthalmologic and cerebellar defects, and ichthyosiform skin lesions [47–49]. The other disorder is Kahrizi syndrome (KHRZ) (MIM: 612713), which has overlapping features with SRD5A3-CDG – it is an autosomal recessive neurodevelopmental disorder characterized by ID, cataracts, coloboma, kyphosis, and coarse facial features [50].

**Table 1.** Mapped autosomal recessive variants for ID. * - affected individual. FID – family ID. IID – individual ID. Genotypes: 0 – reference allele, 1 – alternate allele. DP – depth of sequencing at variant position.

| FID | IID | CHR:POS:REF/ALT<br>(variant type) | Genotype | DP | Intellectual disability | Born |
| --- | --- | --- | --- | --- | --- | --- |
| ARID-76 | IV-1 | 4:55364215: A/G<br>(missense) | 0/1 | 30 | None | 1966-1970 |
|  | V-4* |  | 1/1 | 28 | Moderate | 1991-1995 |
|  | V-1 |  | 0/0 | 42 | None | 2006-2010 |
| ARID-78 | III-1 | 20:56473304: G/A<br>(missense) | 0/1 | 17 | None | 1981-1985 |
|  | III-2 |  | 0/1 | 20 | None | 1981-1985 |
|  | IV-1* |  | 1/1 | 25 | Mild | 2016-2020 |
|  | IV-2* |  | 1/1 | 35 | Mild | 2011-2015 |
|  | III-1 | 14:67692437:C/T<br>(splice donor) | 0/1 | 17 | None | 1981-1985 |
|  | III-2 |  | 0/1 | 20 | None | 1981-1985 |
|  | IV-1* |  | 1/1 | 20 | Mild | 2016-2020 |
|  | IV-2* |  | 1/1 | 48 | Mild | 2011-2015 |
| ARID-79 | II-1 | 5:140796317:C/A<br>(missense) | 0/1 | 26 | None | 1956-1960 |
|  | III-1* |  | 1/1 | 21 | Severe | 1986-1990 |
|  | II-1 | 15:100331016:G/C<br>(missense) | 0/1 | 25 | None | 1956-1960 |
|  | III-1* |  | 1/1 | 39 | Severe | 1986-1990 |
| ARID-80 | III-1 | 9:35403770:C/T<br>(missense) | 0/1 | 57 | None | 1956-1960 |
|  | III-2 |  | 0/1 | 32 | None | 1966-1970 |
|  | IV-1* |  | 1/1 | 29 | Severe | 2001-2005 |
| ARID-81 | IV-1 | 4:121902938: T/A<br>(missense) | 0/1 | 29 | None | 1966-1970 |
|  | V-1* |  | 1/1 | 21 | Severe | 1996-2000 |

**Table 2.**
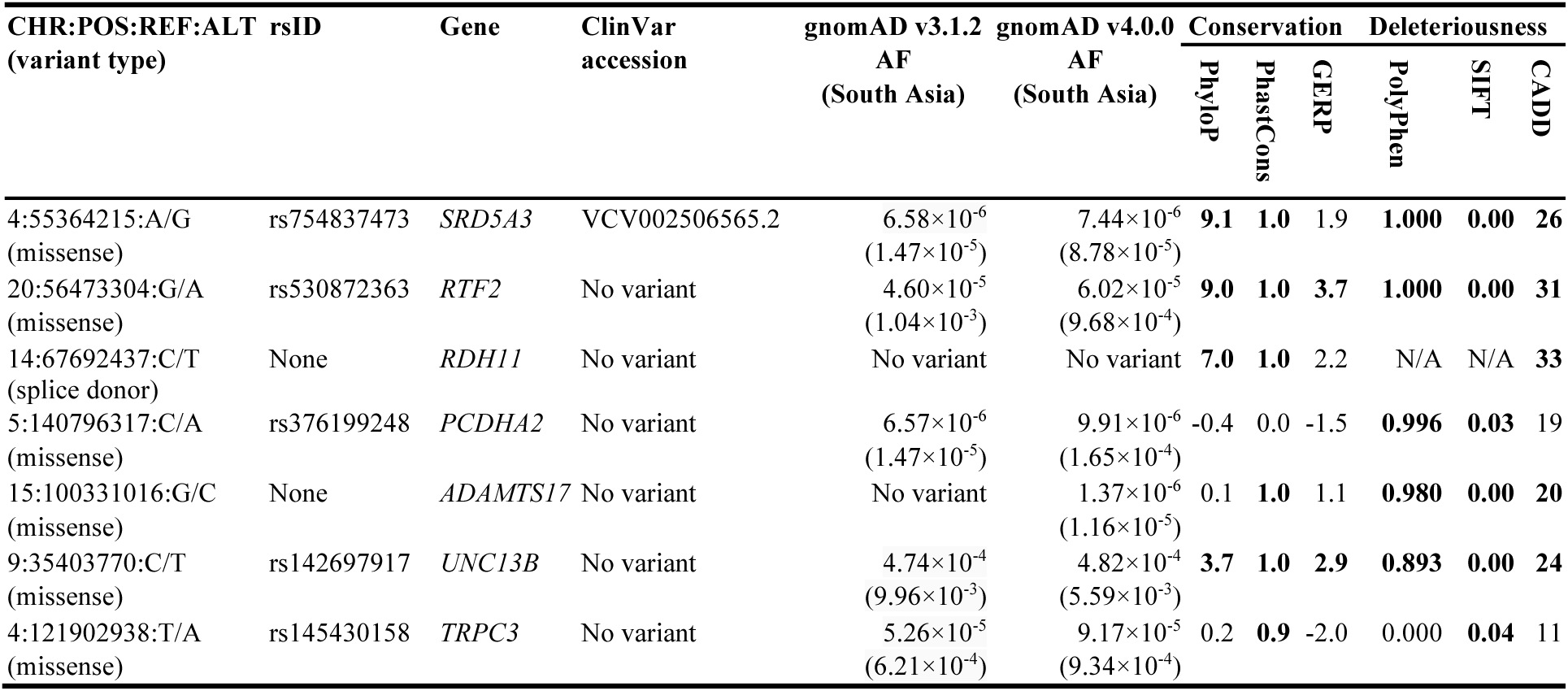
Population frequencies, conservation scores and deleteriousness scores for variants with homozygous alternate genotypes in affected individuals but not in unaffected relatives. Values in bold satisfy the defined thresholds: PhyloP > 1.9, PhastCons > 0.5, GERP > 2.9, PolyPhen > 0.15, SIFT < 0.05, CADD > 20. AF – alternate allele frequency. N/A – not applicable.

To summarize, applying the ACMG guidelines for the variants interpretation [51], we found one moderate and one supportive evidence of pathogenicity. The first evidence, moderate (ACMG PM2), is that the variant is absent from controls or at extremely low frequency if recessive. The second evidence, supportive (ACMG PP3), is that there are multiple lines of computational evidence supporting a deleterious effect. Thus, the variant can be classified as uncertain significance. However, given that the missense variant rs754837473 A>G was already reported in a homozygous state in a carrier with overlapping phenotype, we propose rs754837473 A>G as the most likely cause of ID in the ARID-76 family.

### 3.3. Splice and missense variants inside the *RDH11* and *RTF2* genes in the ARID-78 family

ARID-78 is a four-generation pedigree in which all children of consanguineous parents are affected by ID (Figure 1). WGS was performed on four individuals: III-1, III-2 (unaffected, parents), IV-1 (affected sibling) and IV-2 (affected sibling). Both children show mild intellectual impairment with slightly aggressive behaviour. We found two predicted potentially damaging variants in homozygous states in both affected siblings but not in their parents (Table 1).

The first one was a splice donor variant chr14:67692437 C>T (c.349+1C>T) inside the *RDH11* gene, where both affected individuals had two copies of alternate allele T, while their parents had only one copy. The CADD method predicted the damaging effect of this variant (Table 2), while PolyPhen and SIFT methods were inapplicable to this non-coding variant. The SpliceAI method [52] predicted a splicing donor loss in the canonical transcript with high confidence (delta score = 0.99) due to alternate allele T. The comparative genomic analyses suggested the conservation of the ancestral allele C (PhyloP = 7.0, PhastCons = 1.0, GERP = 2.2), which was present across ten primates (Supplementary Table 6). The variant was not present in ClinVar or gnomAD databases. The number of missense and LoF variants inside the *RDH11* gene reported in gnomAD v4.0.0 is slightly lower than expected, but the difference is not large enough to conclude that the *RDH11* gene may not tolerate missense or LoF mutations (Supplementary Table 5). Several nonsense mutations in the compound heterozygous state in the *RDH11* gene were previously reported as pathogenic in a single family of Italian-American descent, where three siblings had retinal dystrophy, juvenile cataracts, and short stature syndrome (RDJCSS), experiencing psychomotor delays from early childhood, including learning difficulties (MIM: 616108) [53].

The second was a missense variant rs530872363 G>A (p.Asp25Asn) inside the *RTF2* gene. Like the first variant, both affected siblings had two copies of the alternate allele A, while both parents had only one copy. The variant was predicted to be ‘damaging’ by Polyphen, SIFT, and CADD (Table 2). The comparative genomic analyses suggested that the ancestral allele G is conserved across vertebrates (PhyloP = 9.0, PhastCons = 1.0, GERP = 3.7), and it was present across ten primates (Supplementary Table 7). There were no entries for rs530872363 G>A in the ClinVar database. The gnomAD v4.0.0 database had 97 individuals (AF= 6.0×10^-5^) carrying a single copy of the alternate allele A and no homozygous carriers. 88 out of 97 carriers in the gnomAD v4.0.0 database were of South Asian genetic ancestry (AF=1.0×10^-3^), indicating the higher prevalence of this allele in this population. Only seven heterozygous carriers were reported in gnomAD v3.1.2 (no homozygous carriers), and all seven carriers were also present in the gnomAD v3.1.2 non-neuro database. The numbers of missense and LoF mutations observed in the *RTF2* gene in the gnomAD v2.1.1 and v4.0.0 databases are lower than expected. Still, this is only suggestive that this gene may be intolerant to both classes of mutations (Supplementary Table 5). This gene was not known previously for ARID or any other rare condition with a similar phenotype. However, *RTF2* protein levels in the brain were associated with Alzheimer’s disease in recent proteome-wide association studies (PWASs) with mixed evidence of causality [54, 55].

In summary, both chr14:67692437 C>T and rs530872363 G>A have one moderate (ACMG PM2) and one supporting (ACMG PP3) evidence of being pathogenic. Although both variants show evidence of co-segregation with disease in multiple affected family members, none of the genes are known to cause the disease. Thus, the ACMG PP1 criteria for supporting evidence of pathogenicity may not apply. The genotyping of the third affected sibling may help further prioritize one variant over another if one is observed in the heterozygous state. However, lacking additional functional evidence, any remaining variant will still be classified as of uncertain significance.

### 3.4. Two missense variants inside *PCDHA2* and *ADAMTS17* in the ARID-79 family

ARID-79 is a three-generation pedigree where consanguineous parents have two affected and three unaffected children (Figure 1). WGS was done on two family members: the parent (II-1) and the affected child (III-1). The affected child (III-1) has a severe form of ID. They recognize their parents but display aggressive behaviour towards new people. Similarly, their affected sibling (not sequenced) shows profound intellectual impairment with aggressive behaviour. Both affected siblings depend on the family for assistance with daily activities. We found two predicted potentially damaging variants in the homozygous state in the affected child but not in the parent (Table 1).

The first was the missense variant rs376199248 C>A (p.Asn451Lys) in the *PCDHA2* gene. The affected child had two alternate allele A copies, while the unaffected parent had one copy. Only Polyphen and SIFT predicted the variant to be damaging, while the CADD score was just below our threshold (Table 2). Although the ancestral allele C was present across ten primates (Supplementary Table 8), none of the applied algorithms suggested a conservation of it (phyloP=-0.423, phastCons=0.037, GERP=-1.51). The variant was not previously reported in the ClinVar database. The gnomAD v4.0.0 database reported 16 heterozygous alternate allele A carriers (AF=9.91×10^-6^), 13 of non-Finnish European ancestry, and no homozygous. The gene-level constraint scores indicate that the *PCDHA2* gene tolerates missense and LoF mutations (Supplementary Table 5). The *PCDHA2* has not been previously implicated in ARID development. However, it is a part of the *PCDHA* gene cluster, which encodes a family of cadherin-like cell surface proteins expressed in neurons and present at synaptic junctions. Missense mutations in the *PCDHA3* gene were previously reported in patients with restless leg syndrome (RLS), which affects the nervous system and muscles [56]. Moreover, multiple CNVs were reported in patients with developmental delay (DD)/ID and ASD in a region containing the *PCDHA* gene cluster [57, 58]

The second was a missense variant chr15:100331016 G>C (p.Ile163Met) mapped to *ADAMTS17*. The affected child had two alternate allele C copies, while the parent carried only one copy. The Polyphen, SIFT, and CADD methods predicted this variant to be damaging. The ancestral allele G was predicted to belong to a conserved region (phastCons = 0.994) and was present across ten primates (Supplementary Table 9). Still, the remaining methods (GERP and PhyloP) did not indicate conservation at the level of individual nucleotides. The variant was not reported in the ClinVar database, and only two alleles in heterozygous carriers of South Asian and non-Finnish European genetic ancestries were reported in gnomAD v4.0.0. The gene constraint metrics reported in the gnomAD databases indicate that *ADAMTS17* tolerates missense and LoF mutations (only gnomAD v2.1.1 reported a lower number of LoF mutations than expected, but this difference was insufficient to claim for LoF intolerance) (Supplementary Table 5). Mutations inside *ADAMTS17* were not reported for ARID or related phenotypes. Splice-site, frameshift, and truncating homozygous mutations were found in the *ADAMTS17* gene in Indian [59] and Saudi Arabian [60] families with the Weill-Marchesani syndrome 4 (WMS 4) (MIM: 613195), a connective tissue disorder. *ADAMTS17* was highly expressed in the adult lung, brain, eye, and retina in Saudi Arabian families. It’s been reported that 10-17% of the WMS cases may display mild ID [61].

Similar to the ARID-78 family, both identified rare variants in ARID-79 have one moderate (ACMG PM2) and one supporting (ACMG PP3) evidence of pathogenicity and can be classified as of uncertain significance. Although the genotyping of the second affected and the other three unaffected siblings may help eliminate one of the variants, it will not allow us to claim pathogenicity with certainty.

### 3.5. The p. Pro1505Ser mutation inside the *UNC13B* gene in the ARID-80 family

ARID-80 is a four-generation pedigree in which consanguineous parents have two affected and five unaffected children (Figure 1). WGS was performed on both unaffected parents (III-1 and III-2) and one affected child (IV-1). The affected child (IV-1) shows severe intellectual impairment and aggressive behaviour toward others. They need assistance with daily living activities. The other affected sibling (not sequenced) started showing mild intellectual impairment in their 20s, whereas the proband started displaying intellectual impairment during their first several years.

We didn’t find any predicted potentially damaging variants with population-level AF below 0.001% in this family. However, we found one predicted potentially damaging variant after relaxing our AF threshold to 0.01%. The sequenced affected child had two copies of the alternate allele at the missense variant rs142697917 C>T (p.Pro1505Ser) inside the *UNC13B* gene, while both parents had only one copy (Table 1). The Polyphen, SIFT, and CADD methods predicted this variant to be damaging. All three algorithms, phyloP, phastCons, and GERP, suggested that ancestral allele C is conserved across vertebrates and present across ten primates (Supplementary Table 10). The variant was not reported in ClinVar. The gnomAD v4.0.0 database reported 778 carriers of the alternate allele (AF=4.8×10^-4^), including four homozygous carriers. Most carriers, 509 (including all homozygote carriers), were of South Asian genetic ancestry (AF=5.59×10^-3^), indicating the higher prevalence of this variant in this population. The one homozygote carrier was present in the gnomAD v3.1.2 database of controls. The gnomAD databases reported depletion of LoF and missense mutations in *UNC13B* relative to the expected numbers, which may indicate some degree of gene intolerance towards these mutations. (Supplementary Table 5). In general, mutations within the *UNC13B* gene have not been previously associated with ARID. However, *UNC13B* is known to be expressed in the brain (predominantly in the cerebral cortex), and rare nonsense, splice, and missense mutations (in compound heterozygous and heterozygous states) in the gene were reported in Chinese families with partial epilepsy [62]. Multiple CNVs were reported in patients with DD/ID and ASD in a region containing the *UNC13B* gene [57, 58].

Taken together, although the rs142697917 C>T has one supporting evidence of being pathogenic based on computational evidence (ACMG PP3) and may still be viewed as of extremely low frequency in other databases (i.e. satisfy the moderate evidence of pathogenicity ACMG PM2), it also has one strong evidence of being benign. It is observed in healthy adult individuals in the homozygous state (ACMG BS2), while we expect full penetrance at an early age. Despite this, rs142697917 C>T can still be classified as of uncertain significance. Further genotyping of other siblings may be needed to resolve the conflicting evidence of a benign effect with certainty.

### 3.6. The p.Ile793Phe mutation inside the *TRPC3* gene in the ARID-81 family

ARID-81 is a five-generation pedigree in which consanguineous parents have four affected (one deceased) and one unaffected child (Figure 1). WGS was performed on two family members: the unaffected parent (IV-1) and the affected child (V-1). The affected child (V-1) shows severe intellectual impairment, is non-verbal, and entirely depends on their parents for essential self-care activities.

We found only one predicted potentially damaging variant in the homozygous state in the sequenced affected child but not in the parent. The affected child had two copies of the alternate allele A at the missense variant rs145430158 T>A (p.Ile793Phe) inside the *TRPC3* gene, while the parent had only one copy (Table 1). Only SIFT predicted the damaging effect of this variant (Table 2). In contrast, the PolyPhen method predicted benign effects, and the CADD score for this variant was 11, below our set threshold of 20. The phastCons method estimated that the ancestral allele T belongs to the conserved element with a probability of 0.863, and the ancestral allele was present across ten primates (Supplementary Table 11). Other computational methods assessing individual nucleotides didn’t suggest conservation. The variant was not reported in ClinVar. The gnomAD v4.0.0 database reported 148 heterozygote carriers (no homozygous) of the alternate allele (AF= 9.2×10^-5^): 53 heterozygotes were of non-Finish European genetic ancestry (AF = 4.6×10^-5^), and 85 were of South Asian genetic ancestry (AF = 9.3×10^-4^). The gene-level constraint metrics showed suggestive evidence that the *TRPC3* gene is intolerant to LoF mutations and strong evidence that it is intolerant to missense mutations. The region-level constraint metrics also strongly supported intolerance to missense mutations (Supplementary Table 5). The *TRPC3* gene is expressed in the human cerebellum, and the genetic mouse model with the heterozygous gain-of-function mutation in *TRPC3* shows motor and coordination defects associated with the progressive loss of Purkinje cells in the cerebellum [63]. Thus, several studies hypothesized the causal role of this gene in cerebellar ataxia in humans. However, the systematic screening of 108 patients with suspected genetic forms of ataxias failed to reveal any mutations that could significantly contribute to this condition within *TRPC3* [64]. Alternatively, Fogel *et al*., 2014 [65] suggested that the heterozygous missense mutation in a 40-year-old man of European ancestry in the *TRPC3* gene may cause adult-onset spinocerebellar ataxia-41 (SCA41; MIM: 616410), but additional evidence for more robust conclusions were needed.

To summarize, like other discussed mutations, the rs145430158 T>A mutation can be classified as of uncertain significance: it has moderate evidence of pathogenicity because of its low frequency (ACMG PM2); it has supporting evidence of pathogenicity from *in silico* tools (i.e. region-level conservation and deleteriousness as defined by the SIFT score; ACMG PP3). Because the *TRPC3* gene is highly expressed in the cerebellum and may play a role in spinocerebellar ataxia (a neurogenerative disorder), it is possible to speculate that the described mutation may cause ID in the ARID-81 family. The genotyping of other siblings may help resolve this speculation.

### 3.7. The *in silico* modelling predicts changes in *SRD5A3* protein structure caused by the p.Tyr169Cys mutation

We performed *in silico* protein structure modelling for the six reported missense mutations (splice mutation in *RDH11* was excluded from this analysis due to limitations of the applied protein modelling tools) (Table 3). The missense mutation rs754837473 A>G, identified in the ARID-76 family, causes the Tyr169Cys substitution in *SRD5A3* protein-coding transcripts. All four protein structure modelling methods (Venus, FoldX, Rosetta, and Missense3D) predicted the destabilizing effects of the Tyr169Cys mutation on the protein corresponding to the canonical transcript. The Venus, FoldX, and Rosetta methods predicted positive changes in thermodynamic stability, ΔΔG *>* 2 kcal/mol, destabilizing the protein, while Missense3D indicated the cavity alteration in the protein structure. All methods used the in silico AlphaFold predictions as starting structures. The rest of the mutations either didn’t show significant effects on protein structure based on the applied *in silico* methods, or the available *in silico* protein models were not highly accurate. For example, the pLDDT accuracy scores for the available *in silico* protein models for the *UNC13B* gene were below 90, which makes significant positive changes in thermodynamic stability predicted by the FoldX and Rosetta unreliable.

**Table 3.** *In silico* predicted mutation effects on protein folding and structure. Only protein-coding transcripts are reported. The transcript IDs in bold correspond to the Ensembl canonical transcripts. The UniProt IDs in bold correspond to the canonical isoforms. pLDDT – predicted Local Distance Difference test estimates per-residue modelling confidence on the scale from 0 to 100 (lowest to highest). F – submitted compute job failed to complete due to computational/time limits of the web server.

| Gene | Transcript ID | UniProtID | AlphaFoldID (pLDDT) | Mutation | | $\Delta\Delta G$ (kcal/mol) | | | Missense3D |
| --- | --- | --- | --- | --- | --- | --- | --- | --- | --- |
|  |  |  |  | UniProt | AlphaFold | Venus | FoldX | Rosetta |  |
| *SRD5A3* | **ENST00000264228.9** | Q9H8P0 | AF-Q9H8P0-F1 (97.06) | Tyr169Cys | Tyr169Cys | 2.2 | 3.35 | 4.20 | Cavity altered |
|  | ENST00000679836.1 | A0A7P0TBH6 | AF-A0A7P0TBH6-F1 (89.5) | Tyr169Cys | Tyr169Cys | 1.2 | 2.00 | 2.86 | No damage |
| *RTF2* | **ENST00000357348.10** | Q9BY42 | AF-Q9BY42-F1 (81.93) | Asp25Asn | Asp25Asn | -0.2 | 0.30 | 0.48 | No damage |
|  | ENST0000023939.8 | A0A0A0MQR2 | AF-A0A0A0MQR2-F1 (77.06) | Asp55Asn | Asp55Asn | F | 0.81 | 0.40 | No damage |
|  | ENST00000395881.7 | A2A2L6 | AF-A2A2L6-F1 (73.44) | Asp25Asn | Asp25Asn | F | 0.29 | 0.23 | No damage |
|  | ENST00000449062.1 | A2A2L5 | AF-A2A2L5-F1 (70.44) | Asp55Asn | Asp55Asn | F | 1.21 | -0.25 | No damage |
| *PCDHA2* | **ENST00000526136.2** | **Q9Y5H9-1** | AF-Q9Y5H9-F1 (94.48) | Asn451Lys | Asn451Lys | F | -1.19 | 1.30 | No damage |
|  | ENST00000520672.2 | Q9Y5H9-3 | AF-Q9Y5H9-F1 (94.48) | Asn451Lys | Asn451Lys | F | -1.19 | 1.30 | No damage |
|  | ENST00000378132.2 | Q9Y5H9-2 | AF-Q9Y5H9-F1 (94.48) | Asn451Lys | Asn451Lys | F | -1.19 | 1.30 | No damage |
| *ADAMTS17* | **ENST00000268070.9** | **Q8TE56-1** | AF-Q8TE56-F1 (76.76) | Ile163Met | Ile163Met | F | 0.10 | 1.92 | No damage |
|  | ENST00000568565.2 | H3BRA9 | AF-H3BRA9-F1 (67.81) | Ile163Met | Ile163Met | F | -0.26 | 0.93 | No damage |
|  | ENST00000561355.2 | H3BV94 | Not available | Ile46Met | None | None | None | None | No damage |
| *UNC13B* | **ENST00000635942.2** | A0A1B0GUS7 | Not available | Pro4254Ser | None | None | None | None | No damage |
|  | ENST00000636694.1 | A0A1B0GVW8 | Not available | Pro1904Ser | None | None | None | None | No damage |
|  | ENST00000617908.4 | I6L9J0 | AF-I6L9J0-F1 (80.69) | Pro1111Ser | Pro1111Ser | F | 4.16 | 2.80 | No damage |
|  | ENST00000619578.4 | O14795-2 | AF-O14795-F1 (85.03) | Pro1524Ser | Pro1505Ser | F | 4.17 | 2.13 | No damage |
|  | ENST00000378495.7 | **O14795-1** | AF-O14795-F1 (85.03) | Pro1505Ser | Pro1505Ser | F | 4.17 | 2.13 | No damage |
|  | ENST00000396787.5 | B1AM27 | Not available | Pro1536Ser | None | None | None | None | No damage |
| *TRPC3* | **ENST00000379645.8** | **Q13507-2** | AF-Q13507-F1 (72.02) | Ile793Phe | Ile708Phe | 0.4 | 0.53 | 0.24 | No damage |
|  | ENST00000264811.9 | Q13507-3 | AF-Q13507-F1 (72.02) | Ile720Phe | Ile708Phe | 0.4 | 0.53 | 0.24 | No damage |
|  | ENST00000513531.1 | J3QTB0 | AF-J3QTB0-F1 (65.56) | Ile665Phe | Ile665Phe | 0.3 | 0.13 | 0.26 | No damage |

## 4. DISCUSSION

Seven discussed rare mutations in this work have one supporting (ACMG PP3) and one moderate (ACMG PM2) evidence of pathogenicity. Only one out of seven has conflicting evidence of a benign effect (ACMG BS2). Each of these mutations was unique to a single family and was not observed in other families. Some were observed at higher frequencies in the South Asian population in general. Consequently, each identified mutation was placed in seven genes: *SRD5A3*, *RDH11*, *RTF2*, *PCDHA2*, *ADAMTS17*, *UNC13B*, and *TRPC3*. Only the *SRD5A3* gene was previously linked to ID [47–49] and is included in the approved gene panel for ID (v5.0 from 22 March 2023) available at Genomics England’s PanelApp [5]. The rs754837473 A>G (p.Tyr169Cys) missense mutation inside *SRD5A3* identified in this study was also reported as of uncertain significance by one submitter in ClinVar in an individual with overlapping phenotype. The *in silico* protein structure modelling suggested changes in protein structure caused by this mutation. This evidence strongly suggests that rs754837473 A>G (p.Tyr169Cys) is a likely cause of ID. Despite this, according to the ACMG guidance, more evidence from functional studies may be needed to re-classify it as pathogenic. Three identified genes, *RDH11*, *ADAMTS17*, and *TRPC3*, were not previously implicated in ID but were linked to other rare conditions, which include RDJCSS (MIM: 616108), WMS4 (MIM: 613195), and SCA41 (MIM: 616410), respectively. For the two other genes, *PCDHA2* and *UNC13B,* also not known to cause ID, previous studies reported their high expression in the central nervous system [66] and cerebral cortex [67], respectively. Lastly, two recent studies have reported the association between the increased levels of *RTF2* and the risk of Alzheimer’s disease. Taking this on board, we can state that these six genes, not previously known to cause ID, merit further investigations. Genotyping additional family members, which is one of the subjects in our future work, may help us prioritize *RDH11* vs *RTF2* and *PCDHA2* vs *ADAMTS17*, which are pairs of the genes observed within the same family. However, without functional studies, additional evidence of co-segregation will not be sufficient to re-classify observed mutation from uncertain significance to pathogenic. The findings presented here contribute novel and highly needed information for interpreting rare mutations in genetic testing of ID and provide a starting point for further functional studies of novel candidate mutations and genes.

## 5. CONTRIBUTIONS

A.W. collected samples, performed data analyses and wrote the manuscript. R.E., D.P., and J.S. generated and processed sequencing data. F.L. oversaw the sequencing of samples. Z.M. and A.M. provided help with sample collection and financial support. K.T., R.C.M., and C.B. provided critical feedback on the manuscript. G.A. and A.H.K. supervised the Ph.D. research of A.W. V.M. oversaw the sequencing study, and provided critical feedback on the manuscript. D.T. supervised A.W. in data analyses and wrote the manuscript. All authors read the manuscript.

## 6. ACKNOWLEDGMENTS

A.W. was supported by the Higher Education Commission of Pakistan. The sequencing experiments and analyses of sequencing data in this study were supported by the McGill Canada Excellence Research Chair Program in Genomic Medicine (V.M.). V.M. holds a Canada Excellence Research Chair. Calcul Quebec and the Digital Research Alliance of Canada provided the computational resources for this study. The authors would like to thank Claire Le Moigne from the McGill Canada Excellence Research Chair Program in Genomic Medicine for administrative support.

## 8. DATA AVAILABILITY

Data will be made available on reasonable request, which can be directed to Abdul Waheed and Daniel Taliun.

## SUPPLEMENTARY TABLES AND FIGURES

**Supplementary Table 1.**
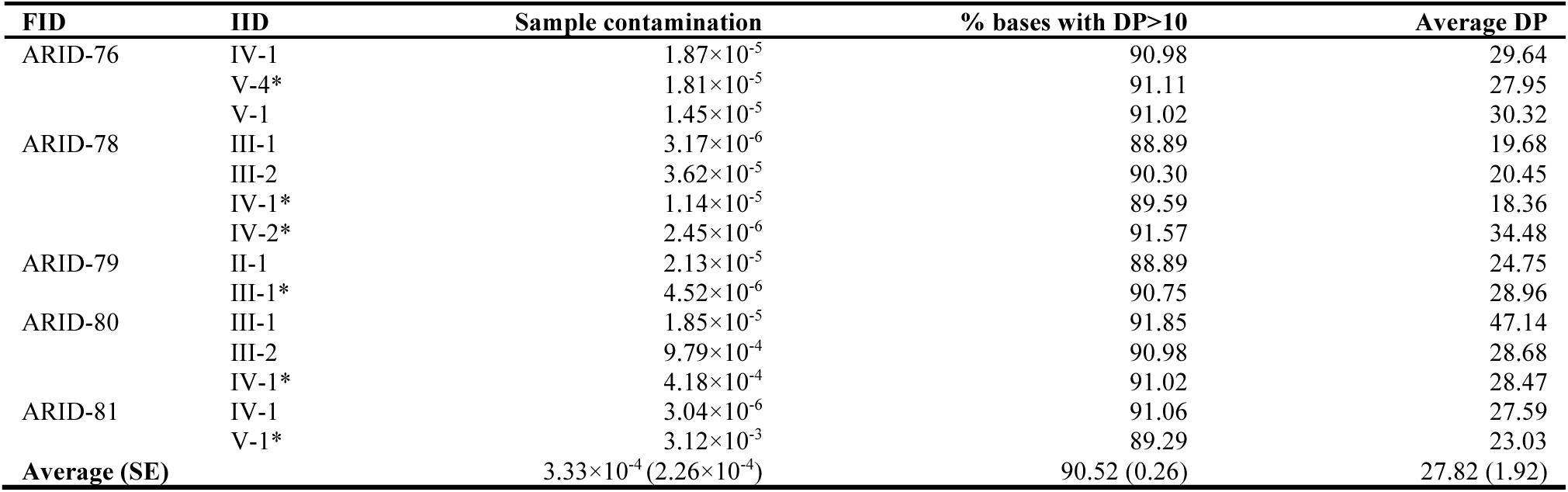
Depth of coverage and estimated contamination rates for individuals in <u>AR</u>ID families. FID – family ID. IID – individual ID. DP – depth of coverage. The sample contamination is a Freemix score computed using VerifyBamId and the HGDP reference panel. * - affected individual.

**Supplementary Table 2.**
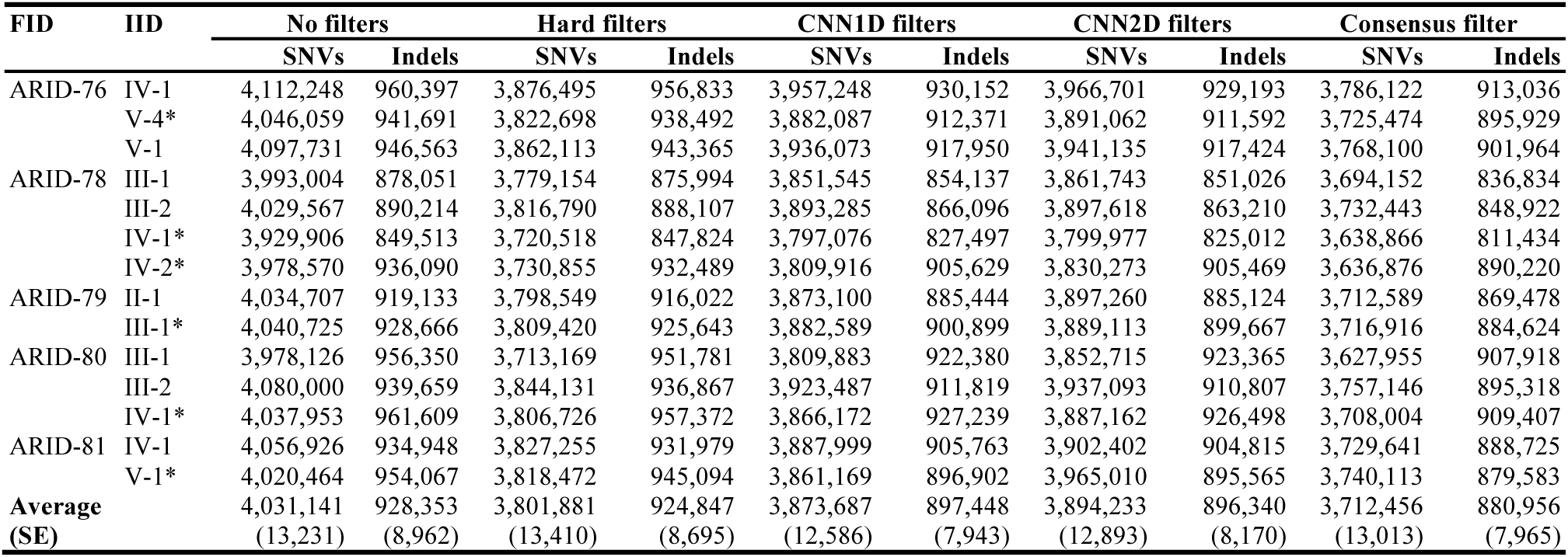
The number of called SNVs and indels per individual. FID – family ID. IID – individual ID. CNN1D and CNN2D are based on Convolutional Neural Network (CNN) scores implemented in GATK CNNScoreVariants. A consensus filter is an intersect of variants which pass all three filters: hard, CNN1D, and CNN2D. * - affected individual.

**Supplementary Table 3.**
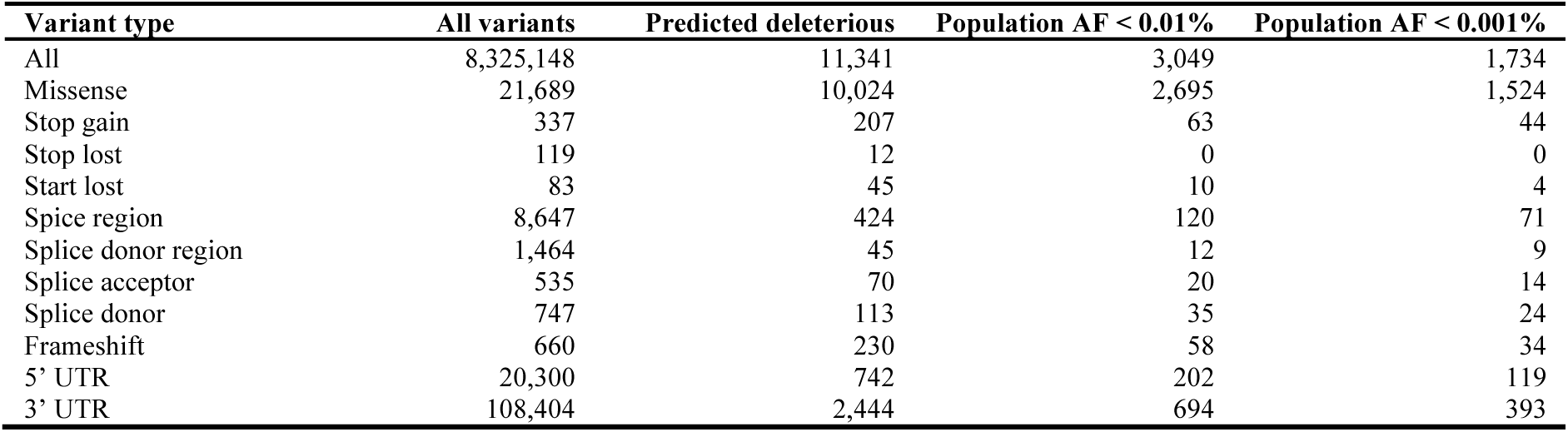
Total number of variants across all sequenced individuals stratified by variant type, deleteriousness, and frequency. When merging variants from multiple individuals, only those variant positions were kept, which had DP > 10 across all individuals. Population alternate allele frequencies are based on gnomAD and 1000 Genomes Project. The variant was predicted deleterious if it had PolyPhen > 0.15 or SIFT < 0.05 or CADD > 20.

| Variant type | All variants | Predicted deleterious | Population AF < 0.01% | Population AF < 0.001% |
| --- | --- | --- | --- | --- |
| All | 8,325,148 | 11,341 | 3,049 | 1,734 |
| Missense | 21,689 | 10,024 | 2,695 | 1,524 |
| Stop gain | 337 | 207 | 63 | 44 |
| Stop lost | 119 | 12 | 0 | 0 |
| Start lost | 83 | 45 | 10 | 4 |
| Splice region | 8,647 | 424 | 120 | 71 |
| Splice donor region | 1,464 | 45 | 12 | 9 |
| Splice acceptor | 535 | 70 | 20 | 14 |
| Splice donor | 747 | 113 | 35 | 24 |
| Frameshift | 660 | 230 | 58 | 34 |
| 5' UTR | 20,300 | 742 | 202 | 119 |
| 3' UTR | 108,404 | 2,444 | 694 | 393 |

**Supplementary Table 4.**
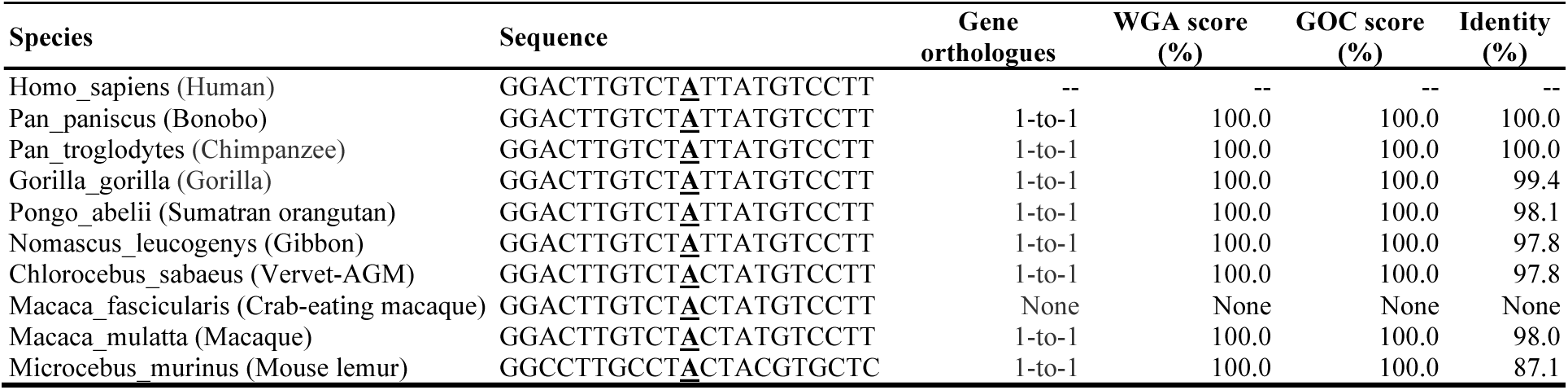
Multiple alignments of 10 primates at 4:55364215:A/G (rs754837473) inside the *SRD5A3* gene. The ancestral allele is marked in bold and underscored. Gene orthologues – 1-to-1(one gene in humans is orthologous to one gene in another species) or 1-to-M (one gene in humans is orthologous to multiple genes in another species).WGA score – Ensembl’s Whole Genome Alignment score between two orthologues. GOC score – Ensembl’s Genome Order Conservation score between two orthologues. Identity – percent of human sequence matching sequence of the orthologue. If human gene has multiple orthologues, then the orthologue with the best WGS, GOC, and identity scores is reported.

**Supplementary Table 5.** Constraint metrics from the gnomAD v2.1.1, gnomAD v4.0.0, and DECIPHER databases. oe – observed vs. expected ratio. LoF – loss of function. CI – confidence interval. The oe LoF, oe missense, and missense Z-score metrics were reported in the gnomAD v2.1.1 database; the missense intolerance score (γ) metric was reported in the DECIPHER database. The missense intolerance score is reported for the region, which overlaps the corresponding mutation within a gene. Values in bold satisfy the recommended intolerance thresholds: missense Z-score >3.09; missense intolerance score (γ) P-value < 0.001.

| Gene | gnomAD v2.1.1 | | | gnomAD v4.0.0 | | | Missense intolerance score ($\gamma$) |
| --- | --- | --- | --- | --- | --- | --- | --- |
|  | oe LoF (90% CI) | oe missense (90% CI) | Missense Z-score | oe LoF (90% CI) | oe missense (90% CI) | Missense Z-score |  |
| *SRD5A3* | 0.54 (0.32 - 0.97) | 0.83 (0.72 - 0.96) | 0.76 | 0.71 (0.52 - 1.00) | 0.91 (0.84 - 1.00) | 0.62 | 0.92 |
| *RDH11* | 0.83 (0.53 - 1.34) | 0.88 (0.78 - 1.01) | 0.55 | 0.59 (0.39 - 0.90) | 0.88 (0.80 - 0.97) | 0.77 | 1.01 |
| *RTF2* | 0.46 (0.27 - 0.84) | 0.72 (0.62 - 0.83) | 1.32 | 0.53 (0.36 - 0.81) | 0.84 (0.76 - 0.93) | 1.06 | None |
| *PCDHA2* | 1.13 (0.85 - 1.51) | 1.09 (1.02 - 1.16) | -0.74 | 0.85 (0.69 - 1.05) | 0.96 (0.92 - 1.01) | 0.47 | None |
| *ADAMTS17* | 0.55 (0.42 - 0.73) | 1.16 (1.09 - 1.23) | -1.39 | 0.83 (0.71 - 0.97) | 1.11 (1.06 - 1.16) | -1.55 | 1.09 |
| *UNC13B* | 0.46 (0.36 - 0.59) | 0.87 (0.82 - 0.92) | 1.45 | 0.64 (0.55 - 0.73) | 0.84 (0.81 - 0.88) | 2.58 | 0.96 |
| *TRPC3* | 0.47 (0.33 - 0.70) | 0.52 (0.47 - 0.58) | **3.84** | 0.59 (0.47 - 0.75) | 0.75 (0.71 - 0.79) | **3.10** | **0.55** |

**Supplementary Table 6.**
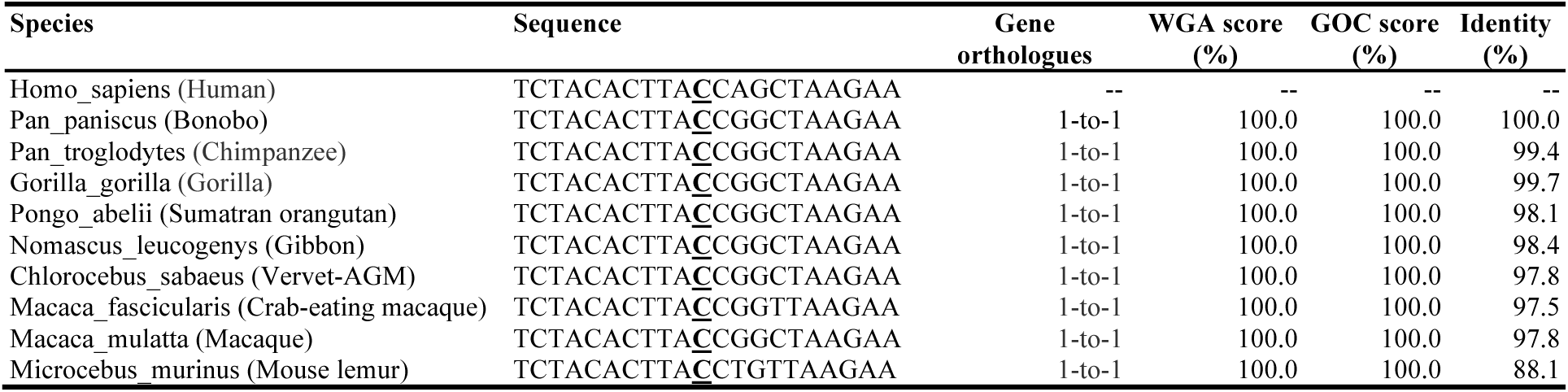
Multiple alignments of 10 primates at 14:67692437:C/T inside the *RDH11* gene. See Supplementary Table 4 for definitions.

**Supplementary Table 7.**
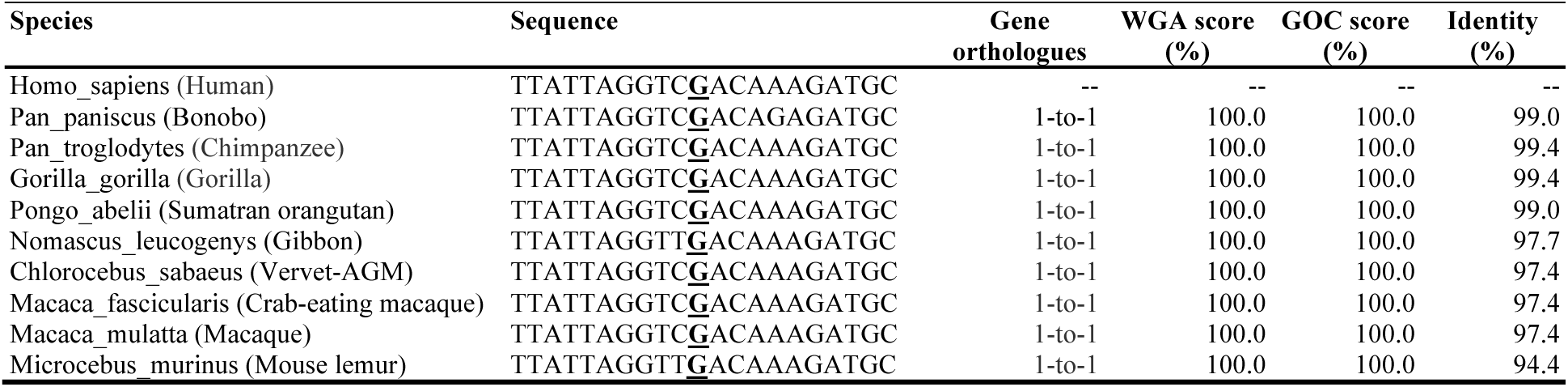
Multiple alignments of 10 primates at 20:56473304: G/A (rs530872363) inside the *RTF2* gene. See Supplementary Table 4 for definitions.

**Supplementary Table 8.**
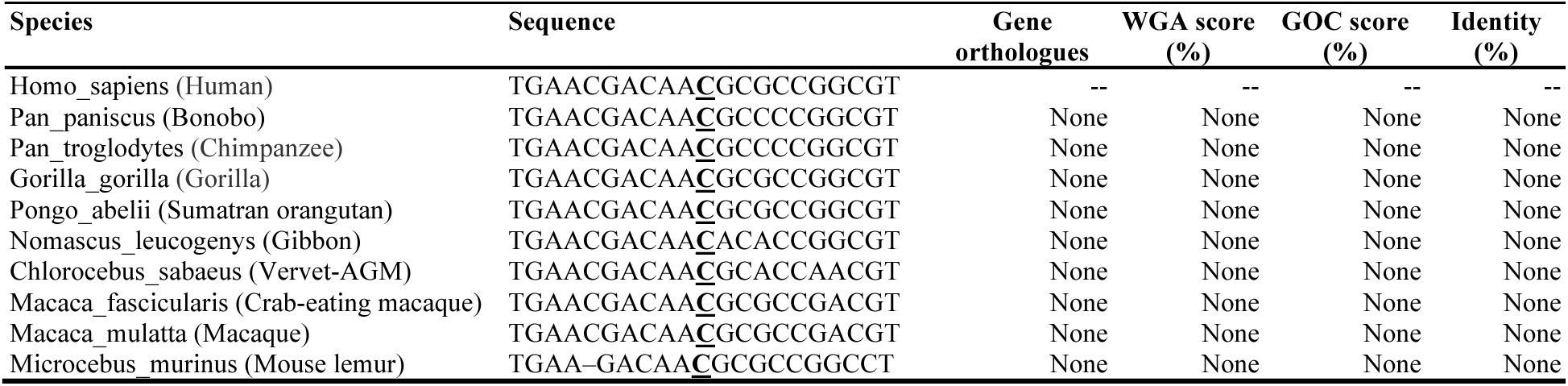
Multiple alignments of 10 primates at 5:140796317:C/A (rs376199248) inside the *PCDHA2* gene. See Supplementary Table 4 for definitions.

**Supplementary Table 9.**
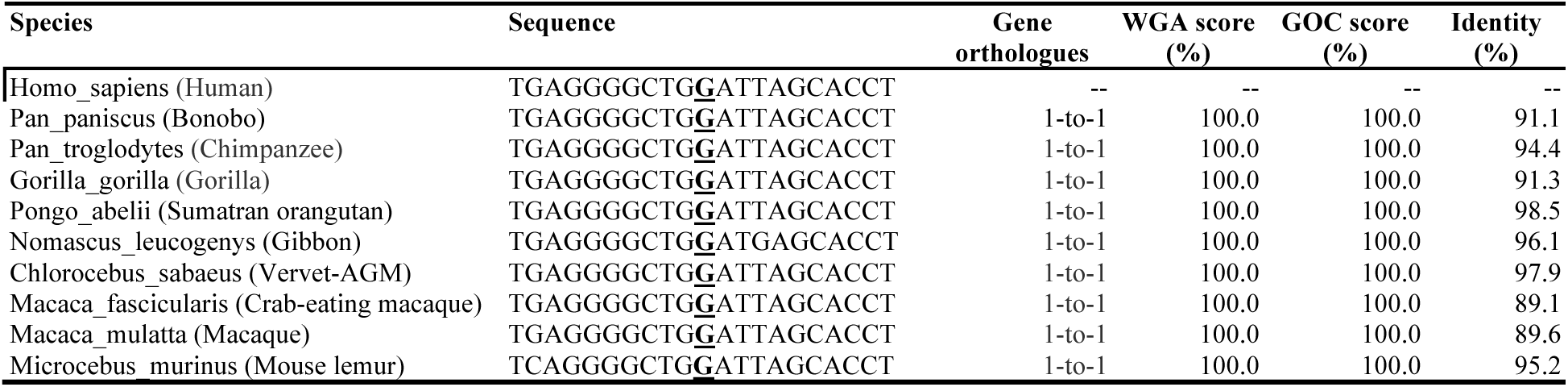
Multiple alignments of 10 primates at 15:100331016:G/C inside the *ADAMTS17* gene. See Supplementary Table 4 for definitions.

**Supplementary Table 10.**
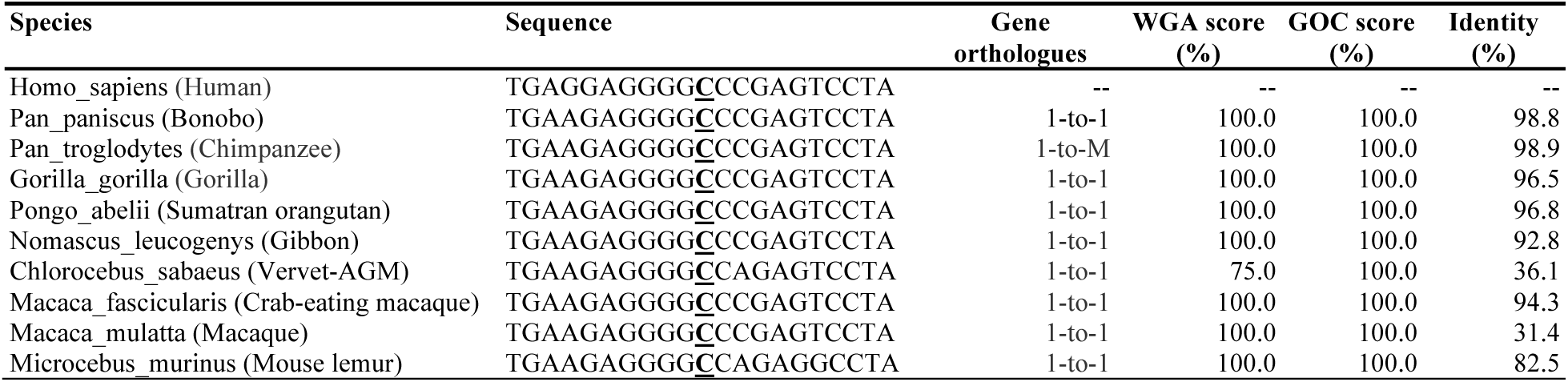
Multiple alignments of 10 primates at 9:35403770:C/T (rs142697917) inside the *UNC13B* gene. See Supplementary Table 4 for definitions.

**Supplementary Table 11.**
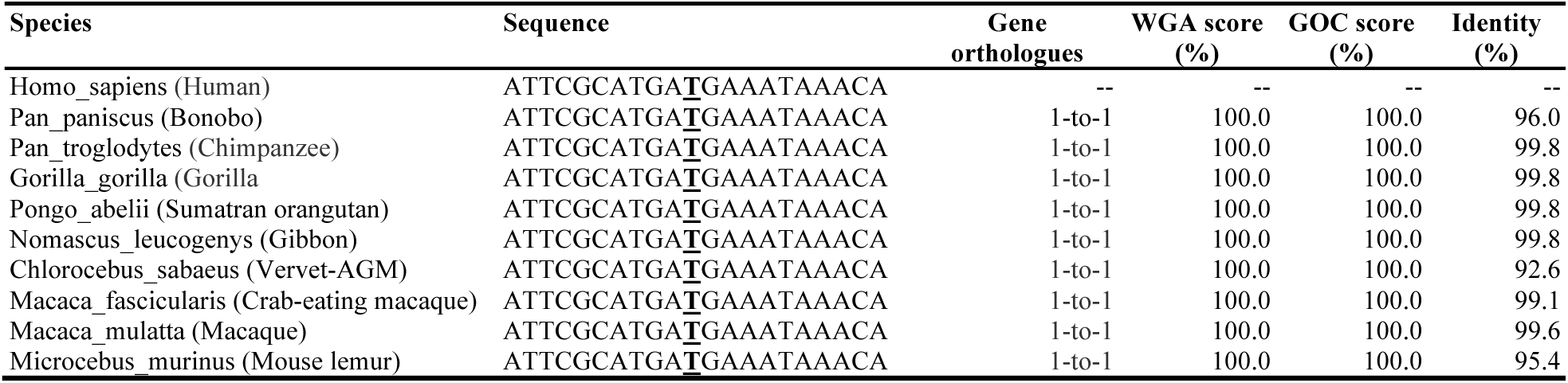
Multiple alignments of 10 primates at 4:121902938: T/A (rs145430158) inside the *TRPC3* gene. See Supplementary Table 4 for definitions.

**Supplementary Figure 1.**
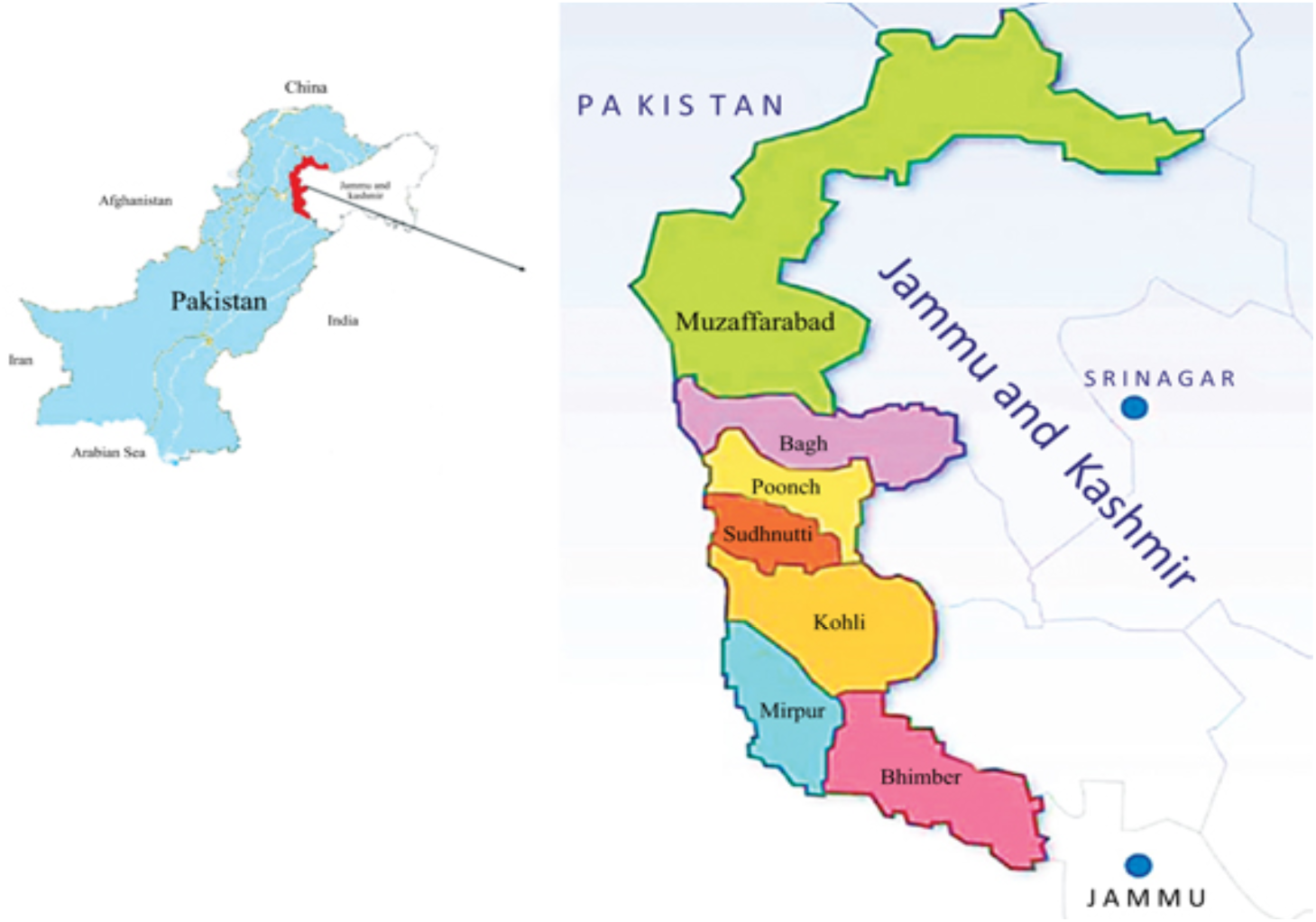
The districts (coloured) of Azad Jammu and Kashmir, Pakistan, where participants were recruited.

**Supplementary Figure 2.**
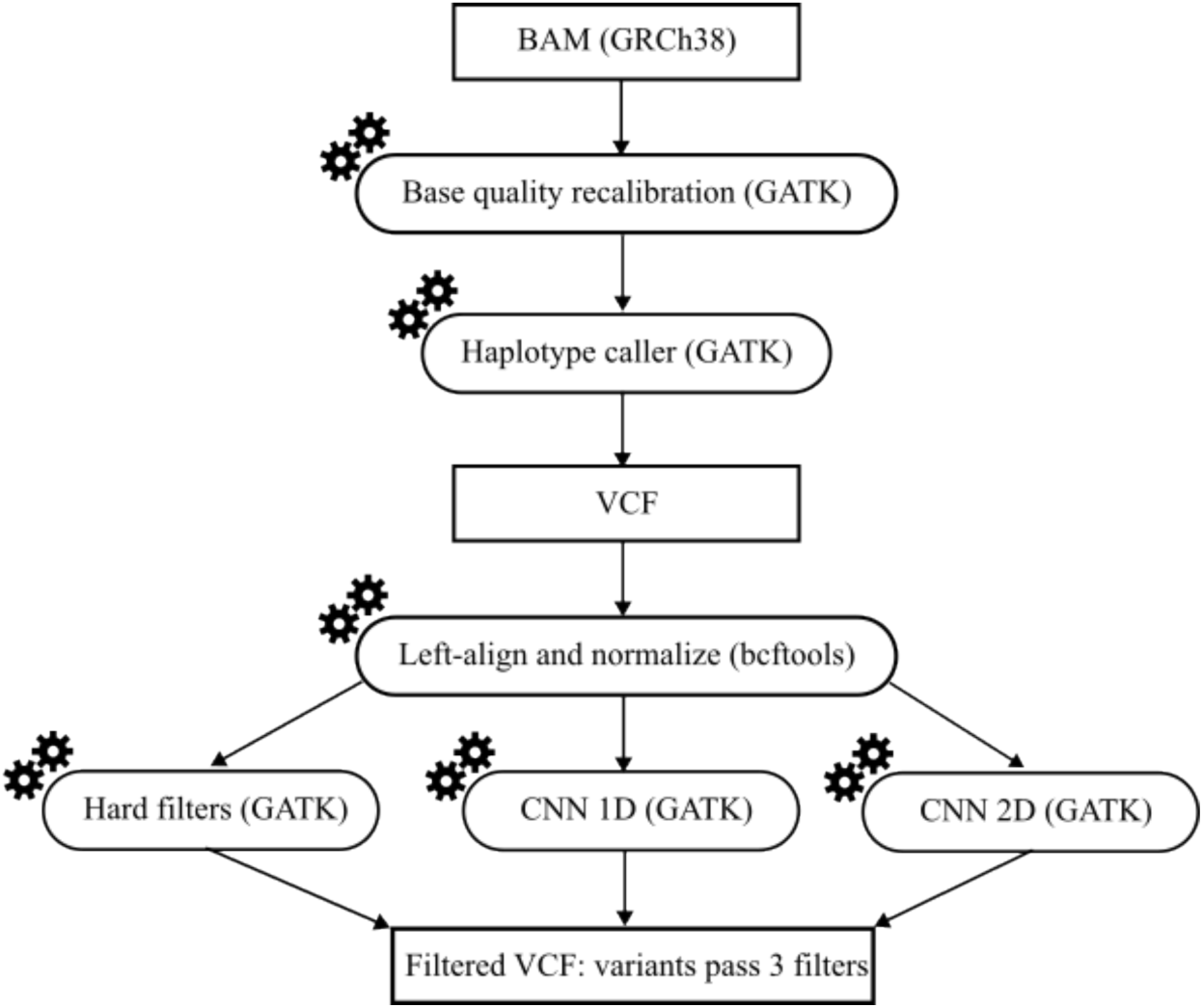
Single sample variant calling and filtering pipeline using GATK.

**Supplementary Figure 3.**
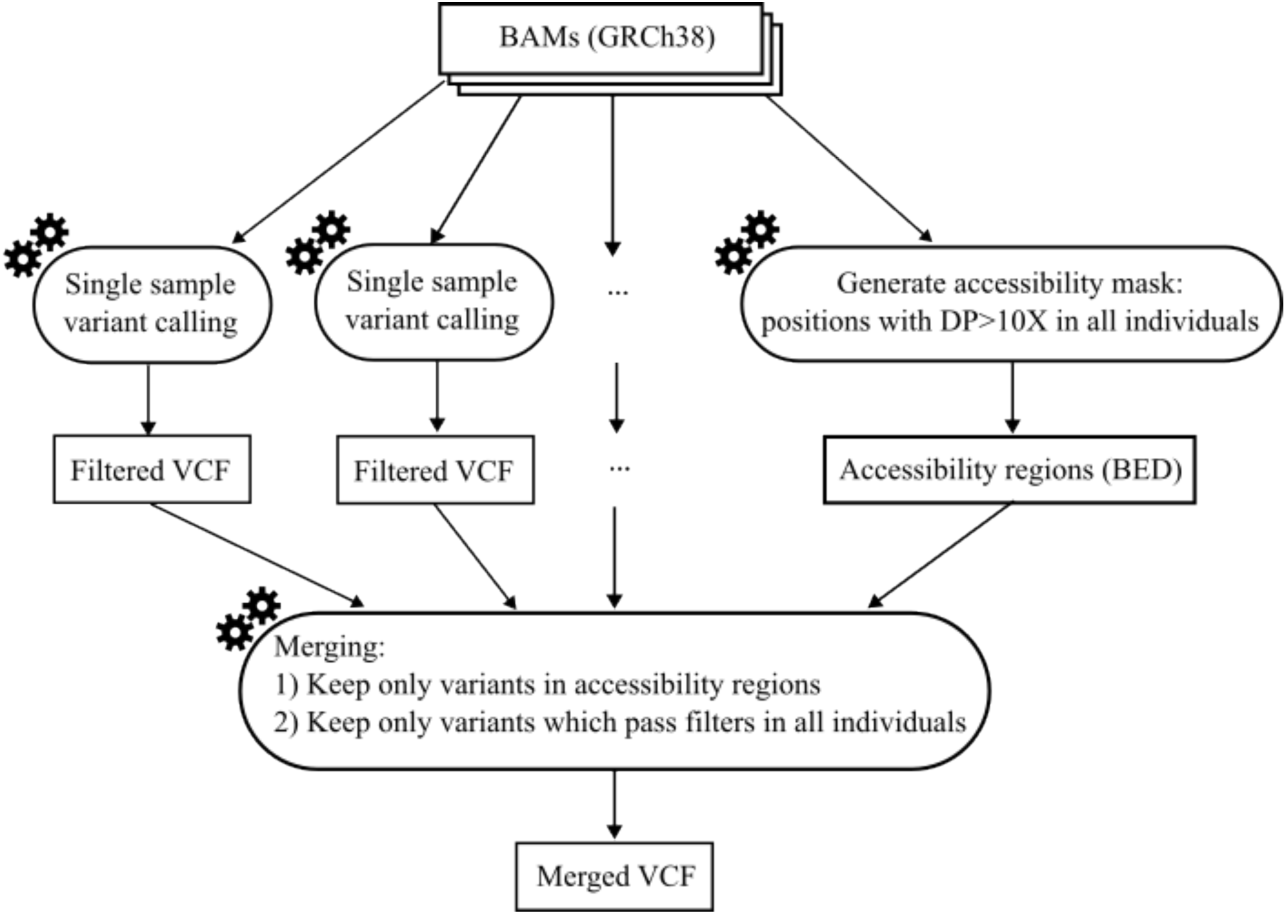
Pipeline for merging single sample VCFs.

**Supplementary Figure 4.**
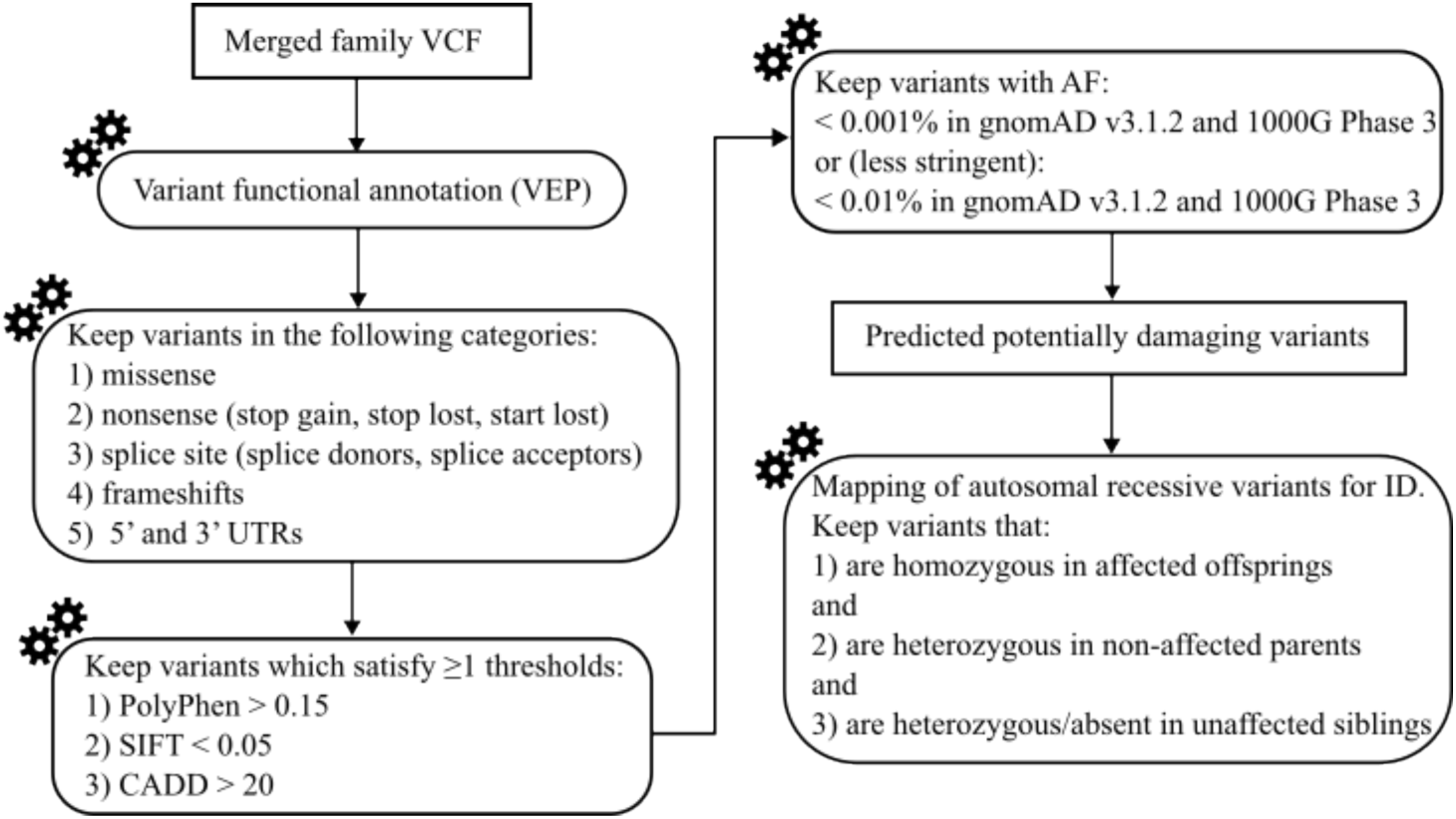
Pipeline for mapping autosomal recessive variants for ID.

**Supplementary Figure 5.**
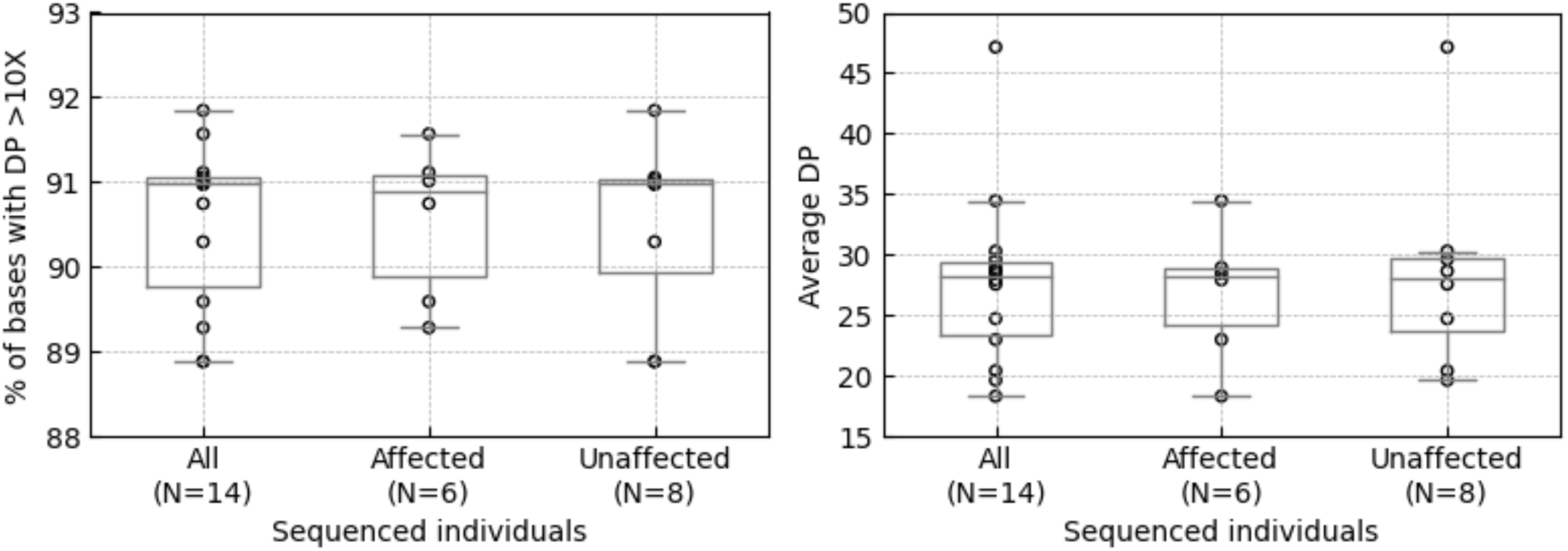
Coverage and depth in sequenced individuals with (affected) and without (unaffected) ID. DP – depths of coverage. The box bounds the IQR, and Tukey-style whiskers extend to a maximum of 1.5 ⨉ IQR beyond the box. The horizontal line within the box indicates the median value. Open circles represent individual data points.

**Supplementary Figure 6.**
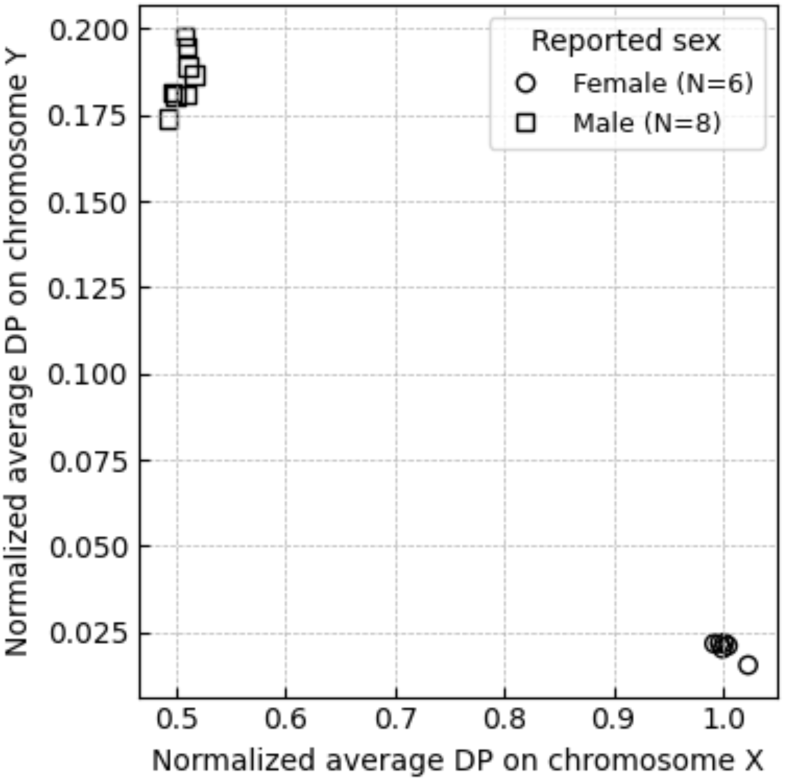
Normalized depth of coverage on sex chromosomes in sequenced individuals. Open circles represent sequenced individuals reported as females. Open squares represent sequenced individuals reported as males.

**Supplementary Figure 7.**
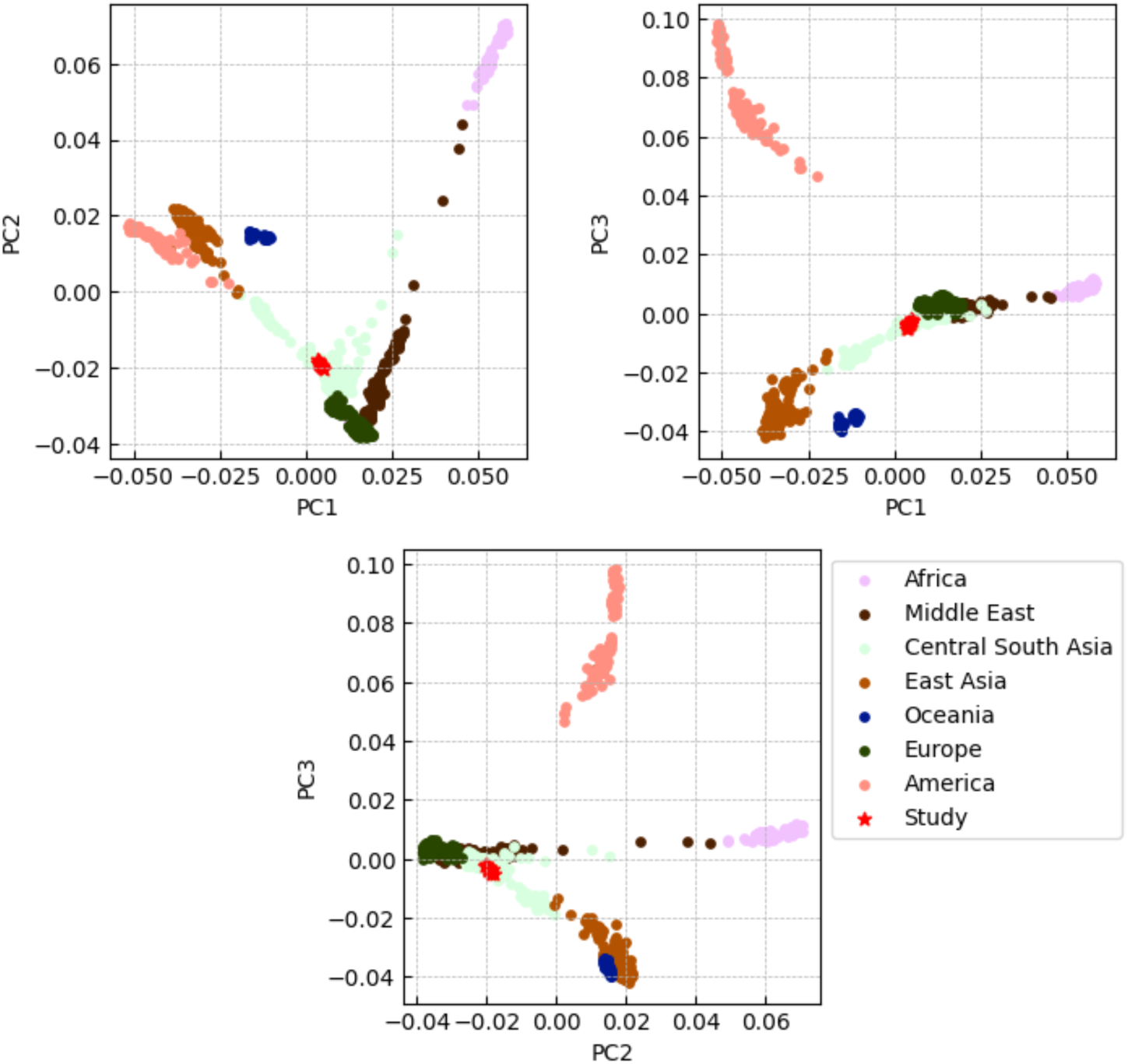
PCA projection of the sequenced individuals into Human Genome Diversity Project (HGDP) reference panel. Red star symbols represent sequenced individuals. Colored circles represent reference individuals from the HGDP dataset.

**Supplementary Figure 8.**
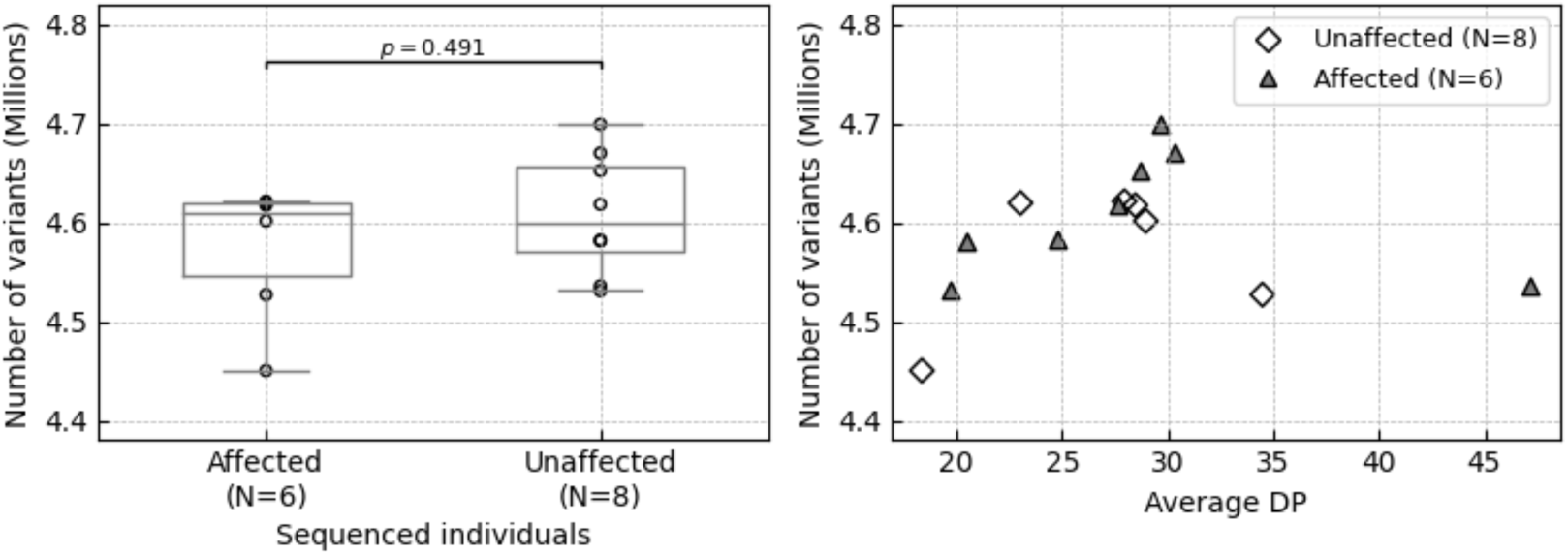
The number of variants in affected and unaffected sequenced individuals. Variants include SNVs and indels, which passed all quality checks. Left panel: the box bounds the IQR, and Tukey-style whiskers extend to a maximum of 1.5 ⨉ IQR beyond the box; the horizontal line within the box indicates the median value; open circles represent individual data points; the p-value corresponds to the two-tailed Mann-Whitney U test. Right panel: DP – depths of coverage; open diamonds represent the number of variants and average sequencing depth in unaffected individuals; shaded triangles represent the number of variants and average sequencing depth in individuals affected with intellectual disability.

